# A Retrospective analysis of DIC score and SIC score in prediction of COVID-19 severity

**DOI:** 10.1101/2021.06.26.21259369

**Authors:** Mayank Kapoor, Prasan Kumar Panda, Lokesh Kumar Saini, Yogesh Arvind Bahurupi

## Abstract

**Background:** The novel Disseminated Intravascular Coagulation (DIC) score [platelet count, prolonged prothrombin time, D-dimer, and fibrinogen] and Sepsis Induced Coagulopathy (SIC) score [platelet count, International normalized ratio, and Sequential organ failure assessment score] are markers of coagulopathy, which, for the first time, are explored in line with the COVID-19 disease outcomes. The correlation of D-dimer with these findings is also studied.

**Patients and methods:** A retrospective analysis of hospital-based records of 168 COVID-19 patients. Data including D-dimer, routine investigations, DIC and SIC scorings (all within three days of admission) were collected and correlated with the outcomes. The study was conducted in a tertiary care center catering to population of North India.

**Results:** Higher DIC score (1·59 ± 1·18 v/s 0·96 ± 1·18), SIC score (1·60 ± 0·89 v/s 0·63 ± 0·99), and D-dimer titers (1321·33 ± 1627·89 v/s 583·66 ± 777·71 ng/ml) were significantly associated with severe COVID-19 disease (P<0·05). DIC score and SIC score ≥ 1, and D-dimer ≥ 1315 ng/ml for severe disease; DIC score ≥ 1, SIC score ≥ 2, and D-dimer ≥ 600 ng/ml for Pulmonary Embolism (PE); and DIC score and SIC score ≥ 1, and D-dimer level ≥ 990 ng/ml for mortality were the respective cut-off values we found from our study.

**Conclusion:** Higher DIC scores, SIC scores, and D-dimer values are associated with severe COVID-19 disease, in-hospital mortality, and PE risk. They can serve as easily accessible early markers of severe disease and prioritize hospital admissions in the presently overburdened scenario, and may be used to develop prognostic prediction models.

**Highlights:** DIC scores, SIC scores, and D-dimer values are hereby studied in association with COVID-19 disease severity, in-hospital mortality, and PE risk. They serve as easily accessible early markers of severe disease and prioritize hospital admissions in the presently overburdened scenario, and may be used to develop prognostic prediction models.

## Introduction

The Coronavirus disease 2019 (COVID-19) pandemic has taken the whole world by storm. Despite the falling number of cases in few areas, it has re-emerged in many countries, raising concerns. Various prognostic markers are proposed to recognize severe disease so that priority is given to those patients first for hospitalization given the over-burdened healthcare infrastructure.

### DIC (Disseminated Intravascular Coagulation) score(1,2)

The International Society for Thrombosis and Hemostasis (ISTH) proposed it as a DIC scoring system. The presence of a disease associated with DIC (malignancy, severe infection, or sepsis) is a prerequisite for using this scoring system. Increasing scores highly correlate with higher mortality rates. The coagulation profile [platelet count, prolonged prothrombin time (PT), D-dimer, and fibrinogen) is a marker of DIC’s dynamic nature (Table 1). These must be repeated serially.

**Table 1:** DIC and SIC scoring system.

| Criteria | DIC Score | SIC Score |
| --- | --- | --- |
| Prolonged PT (seconds) | <3 =0<br>3-5 =1<br>≥6 =2 | N/A |
| INR | N/A | ≤1.2 = 0<br>1.2-1.4 = 1<br>>1.4 = 2 |
| Fibrinogen (mg/dl) | >100 = 0<br><100 = 1 | N/A |
| D-dimer<sup>a</sup> (ng/ml) | No increase = 0<br>Moderate increase = 2<br>Strong increase = 3 | N/A |
| Platelet count (per mm<sup>3</sup>) | >1,00,000 = 0<br>50,000-1,00,000 = 1<br><50,000 = 2 | >1,50,000 = 0<br>1,00,000-1,50,000 = 1<br><1,00,000 = 2 |
| SOFA score | N/A | 1 = 1 point<br>≥2 = 2 points |
| Interpretation | ≥5 points: overt DIC<br><5 points: patient may still have non-overt DIC | Suspect SIC if total score is ≥4. |

### SIC (Sepsis Induced Coagulopathy) score(3)

SIC is the first scoring system that considers the coagulation abnormalities in sepsis following the new Sepsis-3 definition.(4) It is used when the physician considers possible sepsis-induced coagulopathy. It takes into account the Sequential Organ Failure Assessment (SOFA) score values along with the coagulopathy parameters [International Normalized Ratio (INR) and platelet count] (Table 1) and has a high predictive value for 28-days mortality.

D-dimer from the very beginning has been shown to correlate with the severity and mortality in the ongoing COVID-19 pandemic.(5–7) D-dimer forms as a by-product of the clotting mechanism in the body. When a blood clot breaks, D-dimer gets released into the bloodstream. The average D-dimer value is less than 500 ng/ml; any value greater than 500 ng/ml is considered high.(8–12)

As is well known, a patient with COVID infection is predisposed to venous thromboembolism,(13) and this is one of the factors responsible for the worse outcomes. Hence, we explore these new scoring systems, the DIC score and the SIC score, along with the D-dimer values, all of which are markers of coagulopathy, in line with the COVID-19 disease severity, mortality, and the risk of Pulmonary Embolism (PE) development.

## Methods

### Study site

The study was carried out at All India Institute of Medical Sciences, Rishikesh (AIIMS Rishikesh), a tertiary care center catering to North India’s population.

### Participants and Study Design

We retrospectively analyzed all the case records of lab-confirmed COVID-19 patients at least 18 years of age from March 2020 till December 2020. COVID-19 positivity was defined as a positive result on reverse transcriptase-polymerase chain reaction (RT-PCR) SARS-CoV-2 assay of nasopharyngeal or oropharyngeal swab specimens. SD Biosensor standard M nCoV real-time detection kit was used for PCR. Biorad CFX 96 real-time thermocycler, and a Thermo flex 96 extractor machine was used for RNA extraction. All laboratory tests were performed in the institutional laboratory, according to the institutional standards. All patients received routine care as per hospital protocol. The D-dimer levels (calculated by immune-turbidometric assay) and routine blood investigations within three days of hospital admission were collected. DIC scores and SIC scores were calculated according to the respective guidelines.(1),(3) Patients having active thromboembolic disease before symptom onset or before testing for COVID-19 and pregnant females were excluded from the analysis. CT pulmonary angiogram (CTPA) was done based on the clinical judgment of the physician. We categorized the patients during hospitalization into various severity categories according to the Ministry of Health and Family Welfare (MoHFW), India guidelines (Table 2).(14)

**Table 2:** Ministry of Health and Family Welfare (MoHFW) COVID severity categorization(14)

*SpO2: Oxygen saturation*
| Category | Characteristics |
| --- | --- |
| Mild | Without evidence of breathlessness or hypoxia (normal saturation at room air). |
| Moderate | Clinical features of dyspnea, including SpO2 <94% (range 90-94%) on room air, Respiratory Rate ≥ 24 per minute |
| Severe/Critical | Clinical signs of pneumonia plus one of the following; respiratory rate >30 breaths/min, severe respiratory distress, SpO2 <90% on room air |

We determined the correlation of DIC score, SIC score, and D-dimer levels at admission with the clinical outcome and severity in COVID-19 patients and estimated the cut-offs of these parameters in predicting the severity, mortality, and PE risk.

### Sample size

Logistic regression of a binary response variable (Y) on a binary independent variable (X) with a sample size of 127 observations achieved 80% power. Considering a 20 % drop rate, we took a sample size of 168.

### Statistical Analysis

Continuous data following normal distribution and homogeneity of variance was expressed as mean ± SD. Comparison done by independent samples t-test or expressed as median (25–75^th^ percentile) and compared by Wilcoxon rank-sum test. Categorical variables were expressed as number (percentage) and compared by Chi-square tests or Fisher’s exact test. ANOVA followed by Bonferroni’s test was done to correlate the variables with the clinical staging. ROC curves were plotted to derive the cut-offs for various outcomes. Statistical analyses were performed with the SPSS software version 25. P-value < 0·05 was considered statistically significant.

### IEC approval

The Institutional Ethical committee of All India Institute of Medical Sciences Rishikesh approved the study prior to data collection.

**Figure 1:**
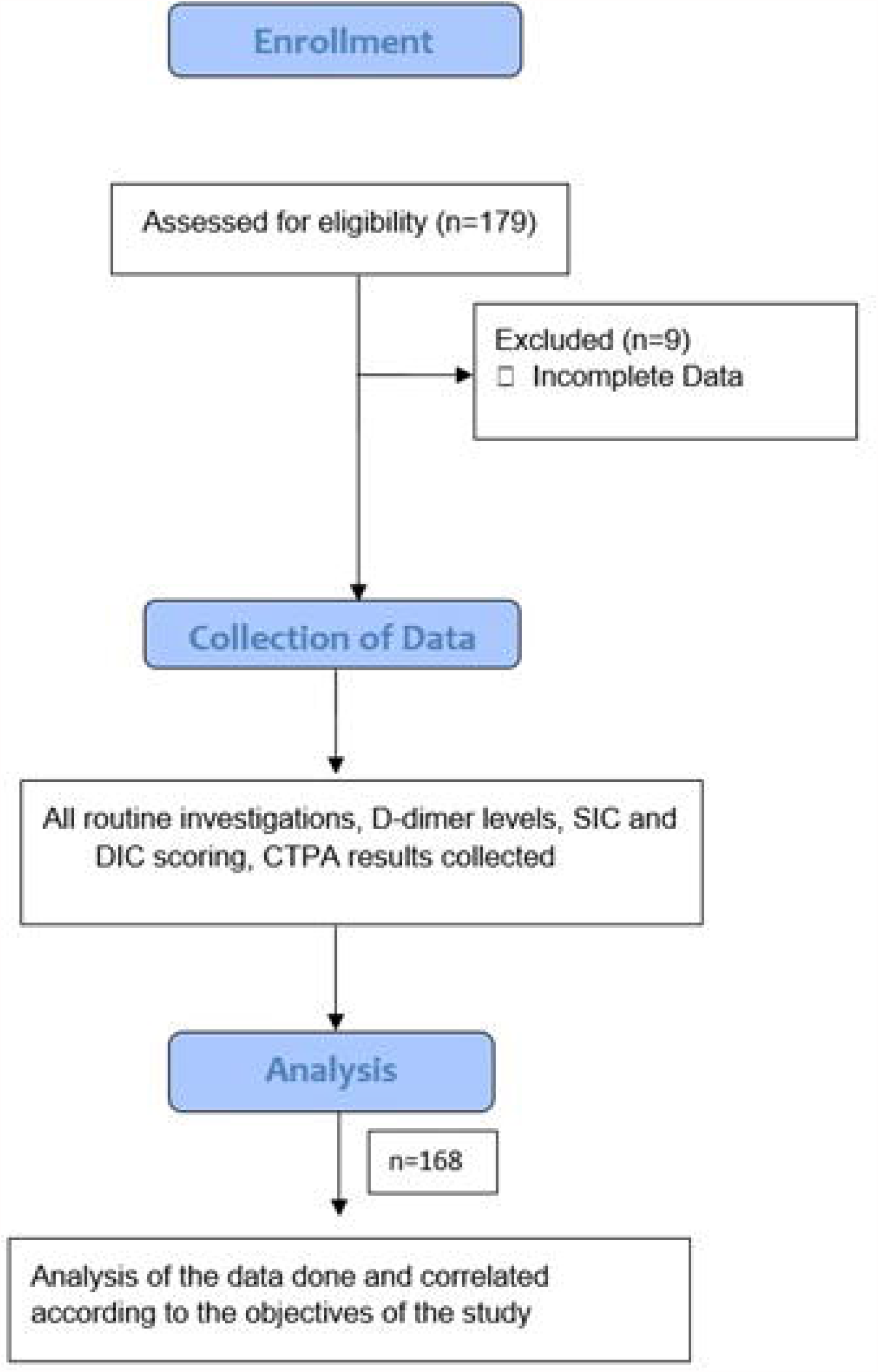
Study Flow

## Results

A total of 168 subjects were recruited in our study (Figure 1). 67·3% (113) of the total patients were males, whereas 32.7% (55) were females. The baseline characteristics of the patients are presented in Table 3. 39·9% (67) of the patients had mild disease, whereas 22·6% (38) had moderate, and 37·5% (63) had severe disease at presentation. The mortality rate was 19·6% (33), while 80·3% (135) patients were discharged from the hospital. Based on the clinician’s recommendation, CTPA was done for 82 subjects. It was reported as normal for 70 (85·3%) patients, whereas PE was detected in 12 (14·6%). Association of the variables with the severity of COVID disease is presented in Table 4. The correlation of DIC scores, SIC scores, and D-dimer values with the various outcomes is presented in Tables 5 and 6. Higher DIC score (1·59 ± 1·18 v/s 0·96 ± 1·18), SIC score (1·60 ± 0·89 v/s 0·63 ± 0·99), and D-dimer titers (1321·33 ± 1627·89 v/s 583·66 ± 777·71 ng/ml) showed a significant correlation with severe COVID-19 disease (P<0·05). Non-survivors also had higher DIC score (2·10 ± 1·22 v/s 1·10 ± 1·29), SIC score (2·00 ± 0·90 v/s 0·89 ± 1·06), and D-dimer levels (2239·16 ± 1825·84 v/s 672·91 ± 972·87 ng/ml) (P<0·05). Similarly, patients with PE demonstrated raised DIC score (2·00 ± 1·59 v/s 1·19 ± 1·27; P=0·05), SIC score (1·92 ± 0·99 v/s 1·26 ± 0·95), and D-dimer titers (2025·80 ± 1814·02 v/s 904·34 ± 1285·98 ng/ml) (P<0·05).

**Table 3:** Baseline Characteristics of the patients.

| Variable | Mean ± SD |
| --- | --- |
| Age (years) | 46.3 ± 15.1 |
| Duration of Hospital stay (days) | 16.3 ± 11.8 |
| **Vitals at the time of hospital admission** |  |
| Temperature (°F) | 98.4 ± 1 |
| RR (per min) | 23.9 ± 5.9 |
| HR (per min) | 91.9 ± 14.9 |
| SBP (mm hg) | 125.1 ± 18.9 |
| DBP (mm hg) | 78.3 ± 11.5 |
| SpO2 (%) | 91.5 ± 7.4 |
| **Lab Parameters** |  |
| TLC (per mm<sup>3</sup>) | 10062 ± 13620 |
| **Hb (gm/dl)** | 11.8 ± 2.4 |
| **D-Dimer (ng/ml)** | 970.9 ± 1326.2 |
| **Fibrinogen (mg/dl)** | 351 ± 242 |
| **Platelet count (thousand/mm<sup>3</sup>)** | 194.1 ± 88.9 |
| **PT (seconds)** | 12.8 ± 2.1 |
| **INR** | 1 ± 0.1 |
| **DIC score** | 1.3 ± 1.2 |
| **SIC score** | 1.1 ± 1 |
| **CTSI** | 23.2 ± 11.5 |

**Table 4:**
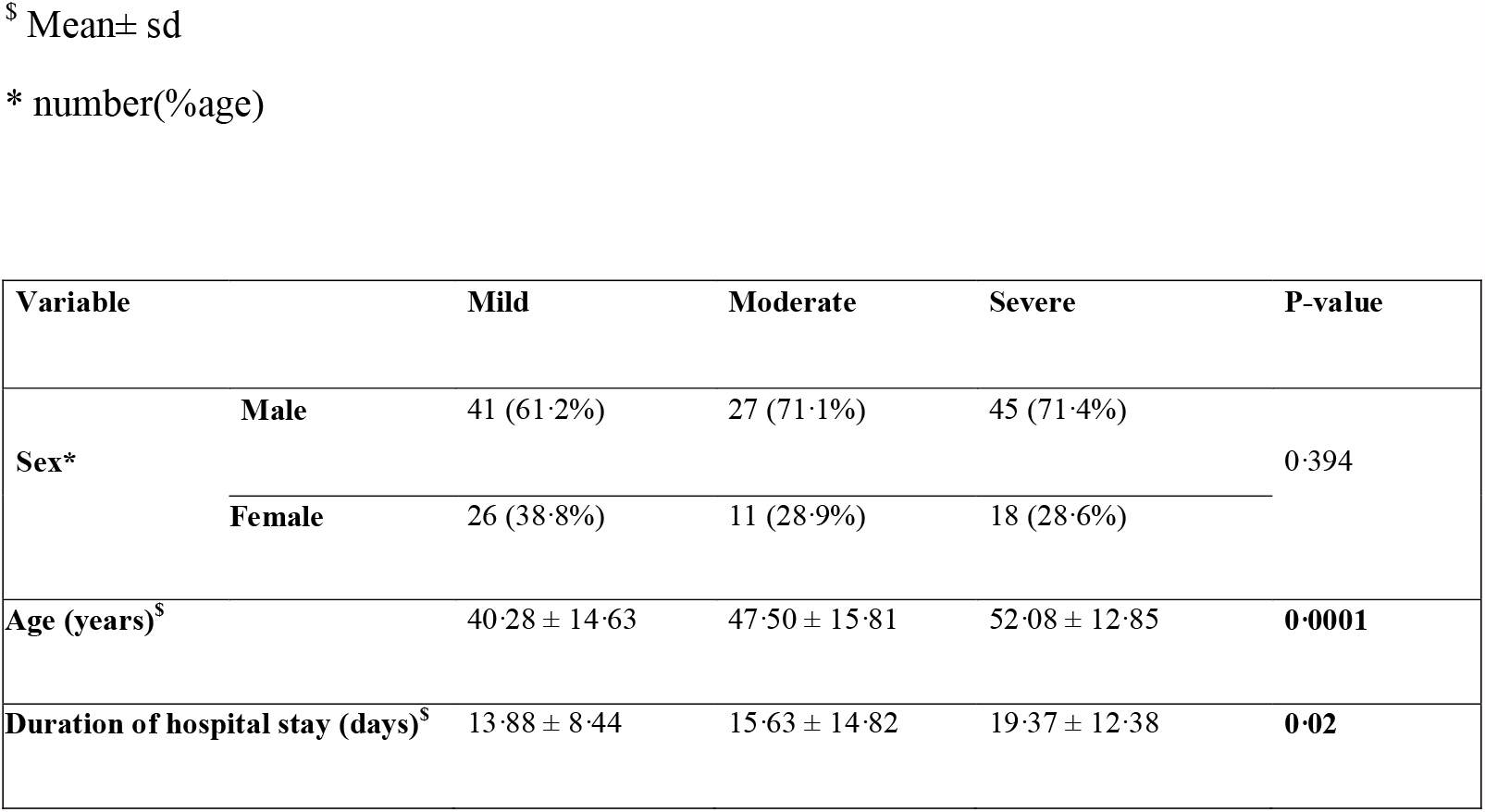
Association of variables with the severity of COVID.

**Table 5:**
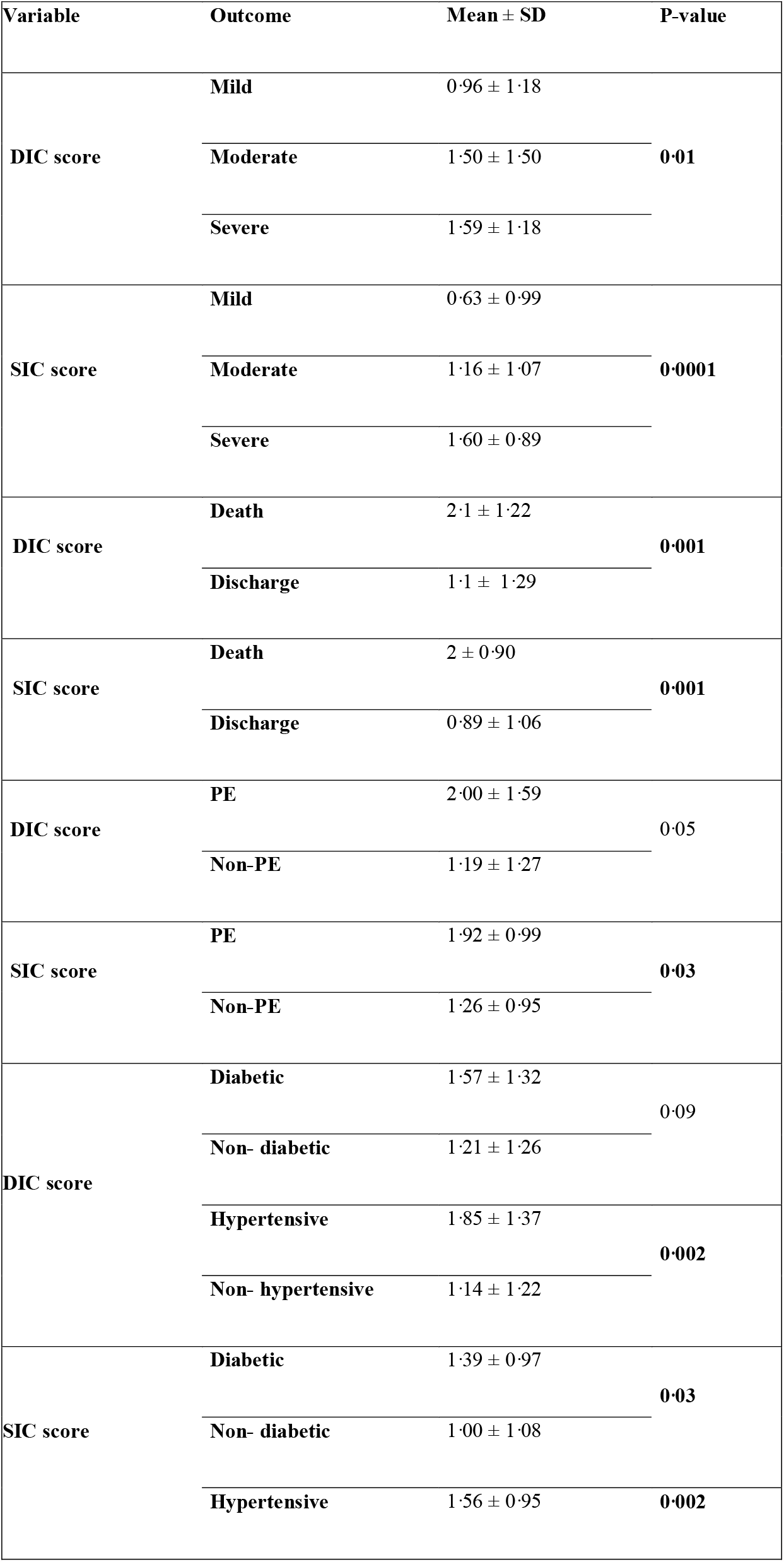
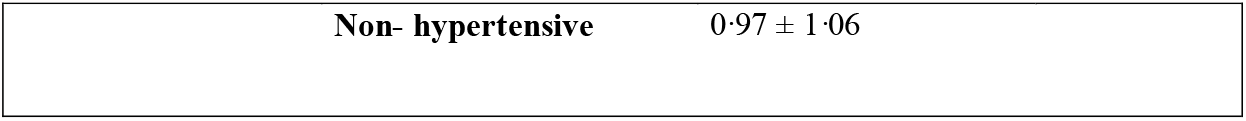
Correlation of DIC scores and SIC scores with outcomes.

**Table 6:** Correlation of D-dimer values with the outcomes.

| Variable | Outcome | Mean ± SD | P-value |
| --- | --- | --- | --- |
| **D-dimer**<br>(ng/ml) | **Discharge** | 672.91 ± 972.87 | **0.0001** |
|  | **Death** | 2239.16 ± 1825.84 |  |
| **D-dimer**<br>(ng/ml) | **Mild** | 583.66 ± 777.71 | **0.005** |
|  | **Moderate** | 1072.82 ± 1388.72 |  |
|  | **Severe** | 1321.33 ± 1627.89 |  |
| **D-dimer**<br>(ng/ml) | **PE** | 2025.80 ± 1814.02 | **0.01** |
|  | **Non-PE** | 904.34 ± 1285.98 |  |
| **D-dimer**<br>(ng/ml) | **Diabetic** | 1281.08 ± 1508.30 | 0.05 |
|  | **Non- diabetic** | 843.22 ± 1227.86 |  |
|  | **Hypertensive** | 1551.19 ± 1686.63 | **0.001** |
|  | **Non- hypertensive** | 783.60 ± 1132.86 |  |

41 patients (24·4%) were hypertensive, whereas 49 (29·16%) were diabetic. Both diabetes mellitus and hypertension were significantly associated with severe disease (Table 7). We analyzed the receiver-operating curve (ROC) cut-offs for the various outcomes. D-dimer showed the highest likelihood ratio (LR+) for all the outcomes followed by SIC score. [Figures 2 (a),(b),(c); Table 8]

**Table 7:** Correlation of Diabetes Mellitus and Hypertension with variables.

| Variable |  | Diabetics | Non-Diabetics | P-value | Hypertension | Non-Hypertensive | P-value |
| --- | --- | --- | --- | --- | --- | --- | --- |
| **PE** | **Yes** | 6 (50%) | 6 (50%) | 0.39 | 3 (25%) | 9 (75%) | 0.72 |
|  | **No** | 26 (37.1%) | 44 (62.9%) |  | 21 (30%) | 49 (70%) |  |
| **Severity** | **Mild** | 5 (7.5%) | 62 (92.5%) | **0.0001** | 5 (7.5%) | 62 (92.5%) | **0.0001** |
|  | **Moderate** | 12 (31.6%) | 26 (68.4%) |  | 9 (23.7%) | 29 (76.3%) |  |
|  | **Severe** | 32 (50.8%) | 31 (49.2%) |  | 27 (42.9%) | 36 (57.1%) |  |

**Table 8:**
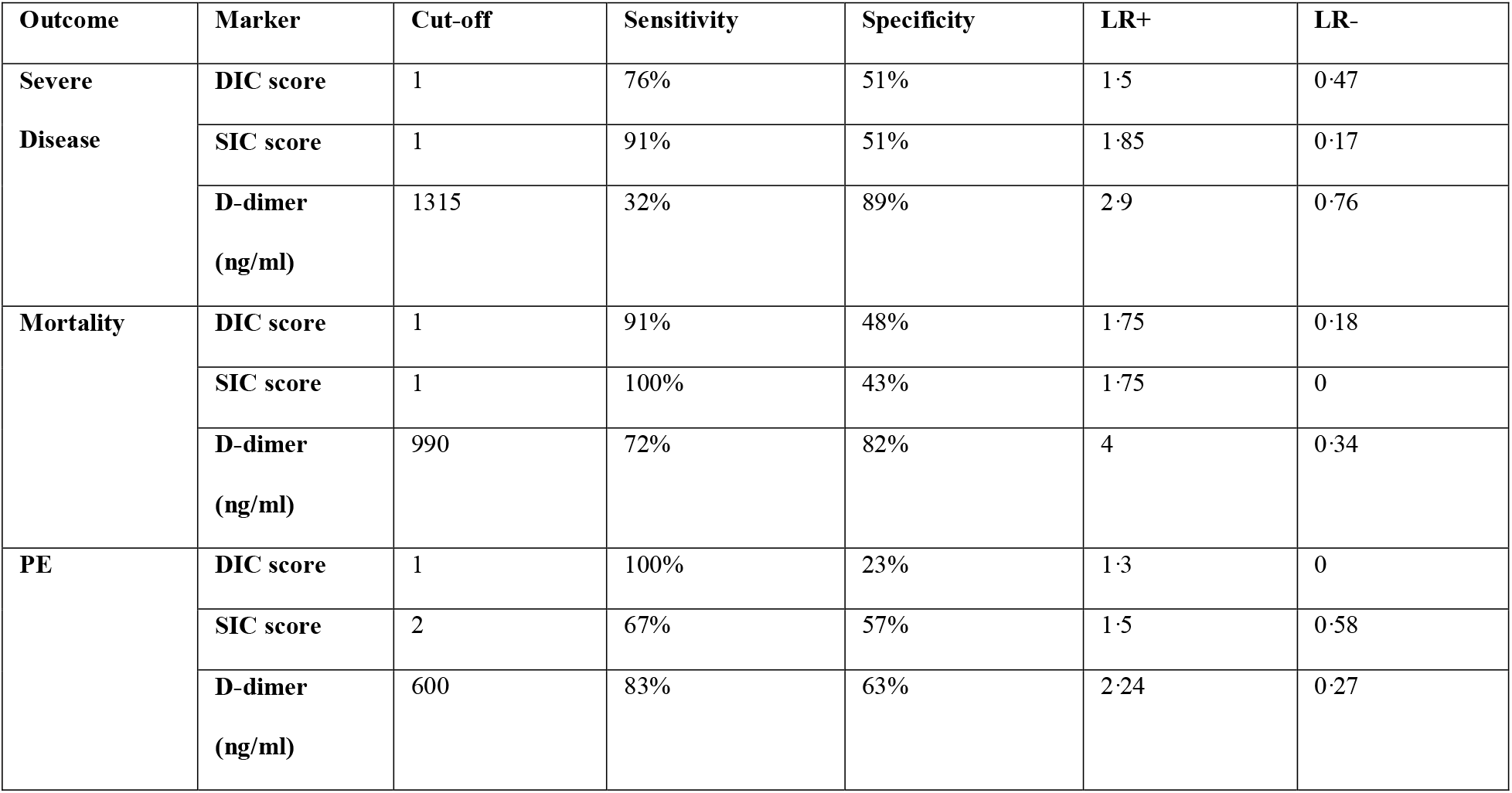
ROC cut-offs of DIC score, SIC score, and D-dimer for severe disease, mortality, and PE.

**Figure 2:**
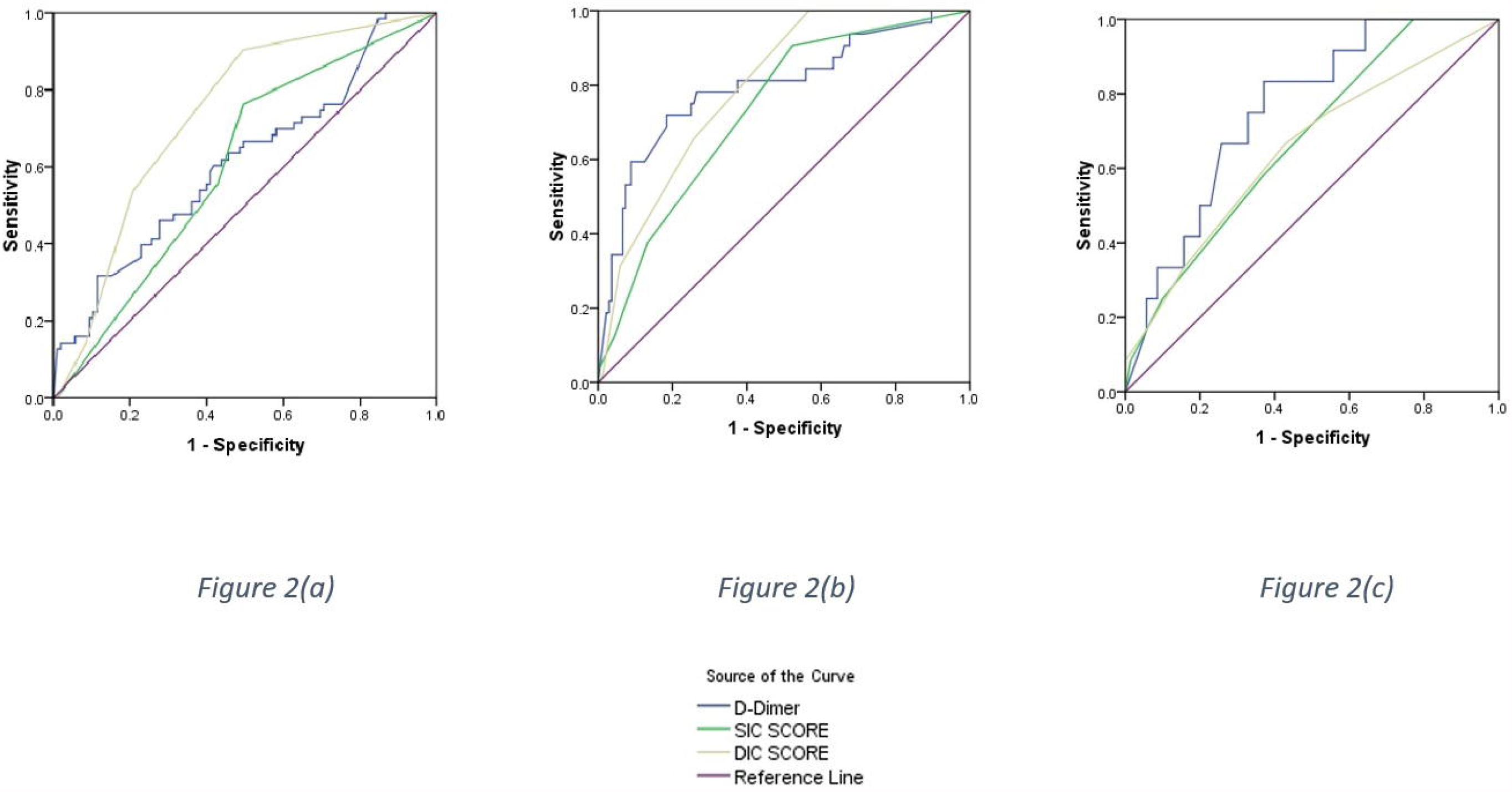
ROC cut-offs (a) Severe disease (b)Mortality (c)PE

## Discussion

The COVID-19 pandemic, although contained in some parts of the world, is still re-emerging in many countries. This has led to the re-introduction of lockdown in some countries.(15) This shows that still, we are far away from altogether tackling this pandemic.(16) It is essential to find out objective parameters to triage COVID-19 patients by predicting severity and outcome; this study was planned with this aim.

To our knowledge, our study is the first of its kind to evaluate the correlation of DIC and SIC scoring with the COVID-19 infection. DIC(1) and SIC(3) scores are scoring systems that predict coagulopathy risk. Any viral infection can progress to sepsis and induce dysfunction in the coagulation system. The COVID-19 infection is unique in having an associated cytokine storm(17) leading to worse prognosis. Integrated analysis has revealed a positive correlation of coagulopathy with cytokine storm in COVID-19 patients. The markers of coagulopathy show a rise early on, indicating that coagulopathy may act as a prodrome of the cytokine storm.(18) Hence utilizing these markers of coagulopathy, we can predict the severity and outcome of the disease. DIC and SIC scores demonstrated

a significant association with the severity of COVID-19 disease in our study, with higher DIC score (1·59 ± 1·18 v/s 0·96 ± 1·18) and SIC score (1·60 ± 0·89 v/s 0·63 ± 0·99) being significantly associated with severe COVID-19 disease (P< 0·05). We propose DIC and SIC score ≥ 1 as the cut-off for predicting severe disease (sensitivity 76%/91%). DIC is a strong predictor of mortality, with studies showing up to 71·4% of the expired patients fulfilling the criteria for DIC compared to 0·6% of the survivors.(19) In our study too, higher DIC (2·1 ± 1·22 v/s 1·1 ± 1·29) and SIC (2·00 ± 0·90 v/s 0·89 ± 1·06) scores demonstrated significant association with mortality (P< 0·05). We found DIC (sensitivity 91%) and SIC (sensitivity 100%) score ≥ 1 as the cut-off for predicting mortality in this study. As is well known, the pro-thrombotic state can lead to events of venous thromboembolism, particularly PE. PE occurs whenever a clot forms within the pulmonary vasculature.(20) SIC scoring showed a significant association with PE probability, whereas DIC scores, although elevated in the PE patients, were not statistically significant. We found DIC (sensitivity 100%) score ≥ 1 and SIC (sensitivity 67%) score ≥ 2 as the cut-offs for predicting PE.

Patients with higher d-dimer values had more severe disease (1321·33 ± 1627·89 v/s 583·66 ± 777·71 ng/ml, P=0·0005). D-dimer’s use as a marker of severity of COVID-19 is under evaluation from the beginning, with various studies stating the same.(21–24) D-dimer levels rise due to the inflammatory reaction associated with COVID, as the inflammatory cytokines released can lead to the imbalance of coagulation and fibrinolysis in the alveolus. This leads to the activation of the fibrinolytic pathway, which increases the D-dimer values.(25),(26) A D-dimer cut-off ≥ 1315 ng/ml predicted severe disease in our study. It has been shown that median D-dimer levels in non-survivors were significantly higher than those in the survivors.(27,28) We also found that non-survivors showed higher D-dimer levels (2239·16 ± 1825·84 v/s 672·91 ± 972·87 ng/ml, P=0·0001) as compared with survivors. Amongst the measured coagulation variables, D-dimer demonstrated the highest ability to predict the mortality in COVID-19 patients in our study, followed by SIC and DIC scores. D-dimer value ≥ 990 ng/mL correlated with higher (sensitivity 72%) in-hospital mortality.(29) COVID patients having PE have demonstrated significantly elevated D-dimer values.(30) Hence, D-dimer forms an excellent marker for predicting PE development in COVID patients; prophylactic anti-coagulation must be considered in patients having higher D-dimer levels to prevent the development of venous thromboembolism. We found higher D-dimer levels in the PE patients (2025·80 ± 1814·02 v/s 904·34 ± 1285·98 ng/ml, P=0·01) as compared to those with normal CTPA findings, and propose a D-dimer cut-off of 600 ng/ml as a threshold (sensitivity 83%) for PE, although further studies are required. Even though previous studies have labeled the cut-off as >2000 ng/ml or even more,(31) our D-dimer value cut-off for PE is lower. One explanation is that we have assessed the D-dimer values within three days of admission and not at the time of PE occurrence, as we are studying D-dimer as a prognostic marker. As is seen with any test, false-negative and false-positive results may be seen. Many physiologic states can lead to elevated D-Dimer levels, including pregnancy, malignancy, smoking, trauma, infections, or sepsis. In addition to these, elderly patients, chronic immobilization, autoimmune diseases, and patients with recent surgeries can also present with high D-Dimer values. An age-adjusted cut-off for D-Dimer is being proposed due to the increase in D-Dimer values as age progresses.(32,33) We can calculate D-dimer values easily, and hence utilizing D-dimer values we can quickly determine patients requiring aggressive care and intensive care unit admissions well in advance.(34)

As most of the patients have diabetes mellitus or hypertension as their significant comorbidities, we evaluated their correlation with the various parameters. Both of these correlated reasonably well with the disease severity. We found the D-dimer levels to be significantly higher amongst the hypertensive (1551·19 ± 1686·63 v/s 783·60 ± 1132·86 ng/ml, P=0·001) as shown in previous studies also.(35) It is well known that people with diabetes are at a higher risk for thrombotic events due to the imbalance between clotting factors and fibrinolysis.(36) Diabetic subgroup has higher D-dimer titers amongst the COVID positive patients.(37) Our study also demonstrated higher D-dimer levels in the diabetics (1281·08 ± 1508·30 v/s 843·22 ± 1227·86 ng/ml, P=0·05). DIC score was high in the hypertensive patients (1·85 ± 1·37 v/s 1·14 ± 1·22), whereas the SIC score was significantly higher in both the hypertensives (1·56 ± 0·95 v/s 0·97 ± 1·06) and the diabetics (1·39 ± 0·97 v/s 1·00 ± 1·08) (For all P<0·05). This again strengthens the fact that both these groups have a higher probability of thromboembolic events, hence demand more focused treatment in the ongoing COVID pandemic.

Our study has limitations. First, owing to our study design we could analyze only the hospitalized patients which allowed us to include fewer mild severity disease cases in comparison to high prevalence of mild COVID 19 cases in the population. So, the results of mild cases cannot be extrapolated to the population as a whole. Hence, larger population studies are needed to confirm our finding. Second, the decision to perform CTPA was purely at the discretion of the treating physician. Hence, CTPA could not be done for all the 168 subjects. As a result, we could have missed some sub-clinical PE cases.

## Conclusion

The novel DIC score, SIC score, and D-dimer levels correlate with the COVID-19 disease severity, in-hospital mortality, and PE probability. Hence, they serve as excellent and easily accessible objective parameters to triage COVID-19 patients by predicting severity and outcome. This can help identify the severe cases early in the disease and decide the bed allocations in the presently overburdened hospitals. We propose the following cut-offs: DIC score and SIC score ≥ 1, and D-dimer ≥ 1315 ng/ml for severe disease; DIC score ≥ 1, SIC score ≥ 2, and D-dimer ≥ 600 ng/ml for PE; and DIC score and SIC score ≥ 1, and D-dimer level ≥ 990 ng/ml for mortality prediction, although further studies are required to strengthen these claims.

## Data Availability

We have access to the data used in the manuscript.

## Acknowledgement

None

